# Polygenic Risk Score Improves Prediction of Primary Open Angle Glaucoma Onset in the Ocular Hypertension Treatment Study

**DOI:** 10.1101/2023.08.15.23294141

**Authors:** Rishabh K. Singh, Yan Zhao, Tobias Elze, John Fingert, Mae Gordon, Michael A. Kass, Yuyang Luo, Louis R. Pasquale, Todd Scheetz, Ayellet V. Segrè, Janey L. Wiggs, Nazlee Zebardast

**Affiliations:** Department of Ophthalmology, Columbia University Medical Center, New York, NY; Schepens Eye Research Institute, Harvard Medical School, Boston, MA; Massachusetts Eye and Ear, Harvard Medical School, Boston, MA; Carver College of Medicine, University of Iowa, Iowa City, IA; Washington University School of Medicine, St. Louis, MO; Icahn School of Medicine at Mount Sinai, New York, NY; Ocular Genomics Institute, Massachusetts Eye and Ear, Boston, MA

**Keywords:** polygenic risk score, genetics, glaucoma, ocular hypertension

## Abstract

**Objective or Purpose:** Primary open-angle glaucoma (POAG) is a highly heritable disease with 127 identified risk loci. Polygenic risks score (PRS) offers a measure of aggregate genetic burden. In this study, we assess whether PRS improves risk stratification in patients with ocular hypertension.

**Design:** A post-hoc analysis of the Ocular Hypertension Treatment Study (OHTS) data.

**Setting, Participants, and/or Controls:** 1636 participants were followed from 1994 to 2020 across 22 sites. The PRS was computed for 1009 OHTS participants using summary statistics from largest cross-ancestry POAG metanalysis with weights trained using 8,813,496 variants from 488,395 participants in the UK Biobank.

**Methods, Interventions, or Testing:** Survival regression analysis, with endpoint as development of POAG, predicted disease onset from PRS incorporating baseline covariates.

**Main Outcomes and Measures:** Outcome measures were hazard ratios for POAG onset. Concordance index and time-dependent AUC were used to compare the predictive performance of multivariable Cox-Proportional Hazards models.

**Results:** Mean PRS was significantly higher for POAG-converters (0.24 ± 0.95) than for non-converters (- 0.12 ± 1.00) (p < 0.01). POAG risk increased 1.36% with each higher PRS decile, with conversion ranging from 9.5% in the lowest PRS decile to 21.8% in the highest decile. Comparison of low-and high-risk PRS tertiles showed a 1.8-fold increase in 20-year POAG risk for participants of European and African ancestries (p<0.01). In the subgroup randomized to delayed treatment, each increase in PRS decile was associated with a 0.52-year decrease in age at diagnosis, (p=0.05). No significant linear relationship between PRS and age at POAG diagnosis was present in the early treatment group. Prediction models significantly improved with the addition of PRS as a covariate (C-index = 0.77) compared to OHTS baseline model (C-index=0.75) (p<0.01). One standard deviation higher PRS conferred a mean hazard ratio of 1.25 (CI=[1.13, 1.44]) for POAG onset.

**Conclusions:** Higher PRS is associated with increased risk for, and earlier development of POAG in patients with ocular hypertension. Early treatment may mitigate the risk from high genetic burden, delaying clinically detectable disease by up to 5.2 years. The inclusion of a PRS improves the prediction of POAG onset.

## Introduction

Glaucoma is the leading cause of irreversible blindness worldwide with primary open angle glaucoma (POAG) accounting for nearly 69% of global prevalence^1^. Early identification of high-risk individuals and therapeutic intervention is necessary to prevent glaucoma-related vision loss. While POAG may present with or without elevated intraocular pressure (IOP)^2^, IOP remains the only modifiable risk factor for its development and progression^3–7^. The Ocular Hypertension Treatment Study determined that early treatment with topical IOP-lowering therapy reduces the cumulative incidence of POAG among patients with ocular hypertension (OHTN)^4^. A prediction model including age, IOP, central corneal thickness (CCT), vertical cup-disc ratio (VCDR), and visual field pattern standard deviation (PSD) was developed to identify participants with the highest baseline risk of developing POAG^4^. This prediction model was confirmed in the European Glaucoma Prevention Study and the Diagnostic Innovations in Glaucoma Study^6,8^.

POAG is a highly heritable disease, with 127 identified common risk variants to date^9,10^. While each variant individually has a small effect, they can be used in the aggregate to measure overall genetic burden with a polygenic risk score (PRS). Genotyping can be done inexpensively, once per lifetime, and enable the calculation of a PRS for a wide range of diseases^11^. PRS may be calculated earlier in life, prior to the impact of environmental or phenotypic risk factors, and recalculated as knowledge of the genetic architecture for disease improves ^11,12^. Since the absolute score for a disease varies with the number of single nucleotide polymorphisms (SNPs) included, PRS is often transformed to a Z-score or percentile^13^. Prior work has shown PRS to be associated with risk and outcomes for a range of complex diseases^14–16^. Genome-wide PRS have been used to effectively identify individuals as high risk for common diseases including coronary artery disease, obesity, atrial fibrillation, and type II diabetes^17,18^. In ophthalmology, PRS have been developed for risk stratification in macular degeneration, diabetic retinopathy, and POAG^19–23^. High POAG PRS has been previously associated with an increased risk of glaucoma, increased risk of development of severe disease, maximum recorded IOP, and younger age at diagnosis^19,20,24^. In patients with existing POAG diagnosis, high PRS was also associated with increased rates of visual field progression and retinal nerve fiber layer thinning^25^. PRS have also been used to enhance prediction models for risk stratification in macular degeneration^26^.

While the OHTS prediction model identified key risk factors for conversion to POAG, more than two-thirds of OHTS participants never progressed from OHTN to POAG. A prior analysis has identified a variant in TMC01 locus (rs4656461) conferring a 12% increase in cumulative POAG incidence in a subgroup of participants^27^. A PRS measures a larger component of genetic burden; however, its relationship to glaucoma risk in patients with OHTN remains unknown. Characterization of underlying genetic risk with PRS has the potential to improve overall POAG risk stratification beyond demographic factors and ocular biomarkers (age, IOP, CCT, VCDR, and PSD) in the OHTS prediction model. The purpose of our study was to use available genetic data from OHTS to understand the relationship between a PRS and POAG risk among OHTN patients, identify associations between PRS and known POAG risk factors, and improve prediction of POAG-onset in OHTS participants using the PRS.

## Methods

### Data collection

This study was a secondary analysis of data from OHTS^4,28,29^. OHTS was a randomized clinical trial assessing the safety and efficacy of early treatment with topical IOP-lowering therapy among patients with ocular hypertension. Details of the OHTS study methods have been reported previously^4,28,29^. Our study was exempt from institutional review board approval as we used only de-identified data. All methods adhered to the tenets of the Declaration of Helsinki for research involving human participants.

In the OHTS, 1636 participants with OHTN were recruited from 22 sites. Prior to randomization, each participant was required to have a Humphrey visual field (HVF) testing and optic nerve head (ONH) photograph, each demonstrating no signs of glaucomatous damage. Randomization of participants to observation or treatment began in 1994 and ended in 1996. Participants were followed with HVFs semi-annually and ONH photography annually until the completion of phase 1 on June 2002. After each visit, the Optic Disc Photography Reading Center assessed serial images masked to the chronological sequence to identify evidence of disc changes. The Endpoint Committee ascertained the cause of changes based on the review of clinical history. OHTS phase 2 began in June 2002 and continued until March 2009. During this phase, all participants, including the phase 1 observation group, received topical IOP-lowering therapy. OHTS phase 3 began in July 2015 and continued until June 2020, assessing the cumulative incidence of POAG over 20 years. Data analysis in this study was limited to 20 years of follow-up per participant.

### OHTS genotyping and analysis

From the 1,636 original participants in OHTS, 1,077 were enrolled in an ancillary genetics study with methodology described in Scheetz et al (2016)^27^. 1,063 participants had DNA samples collected with ^27^ genotyping completed using Illumina Omni-1M-Quad microarrays. As previously reported, quality control steps were applied with GenomeStudio and PLINK 1.07 to remove poor quality samples (call rate < 87.9%), and variants (minor allele frequency (MAF) < 1%, call rate < 99%, Hardy-Weinberg Equilibrium (HWE) < 10e-6) resulting in 1,057 samples and 905,636 variants^27^. The International HapMap Project reference panel was used for imputation. We additionally examined discrepancies between genotype-inferred and self-reported sex and ancestry, as well as cryptic relatedness using identity by descent (IBD; Pi-hat>0.1875 representing second to third cousins or closer). One participant with unresolved differences between genotype-inferred and reported sex was removed. Participants were categorized based on inferred genetic ancestry (Supplement 1). European and African ancestry were most common in OHTS participants and retained for this analysis, after removing two samples with mismatched self-report ancestry and inferred ancestry (N = 1,012) (Supplement 1) (Fig 1). Three out of the remaining 1012 were excluded due to a lack of central corneal thickness data (final N = 1009) (Fig 1). Lastly, all baseline parameters were stratified into participant-level (N=1009) and eye-level (N = 2018) data for statistical analysis. Associations between PRS and baseline ocular parameters and demographics were evaluated with participant-level inputs and outputs. Associations between PRS and time-censored POAG-onset data were evaluated with eye-level inputs and outputs, as 53.20% of POAG conversion in OHTS occurred monocularly.

**Figure 1.**
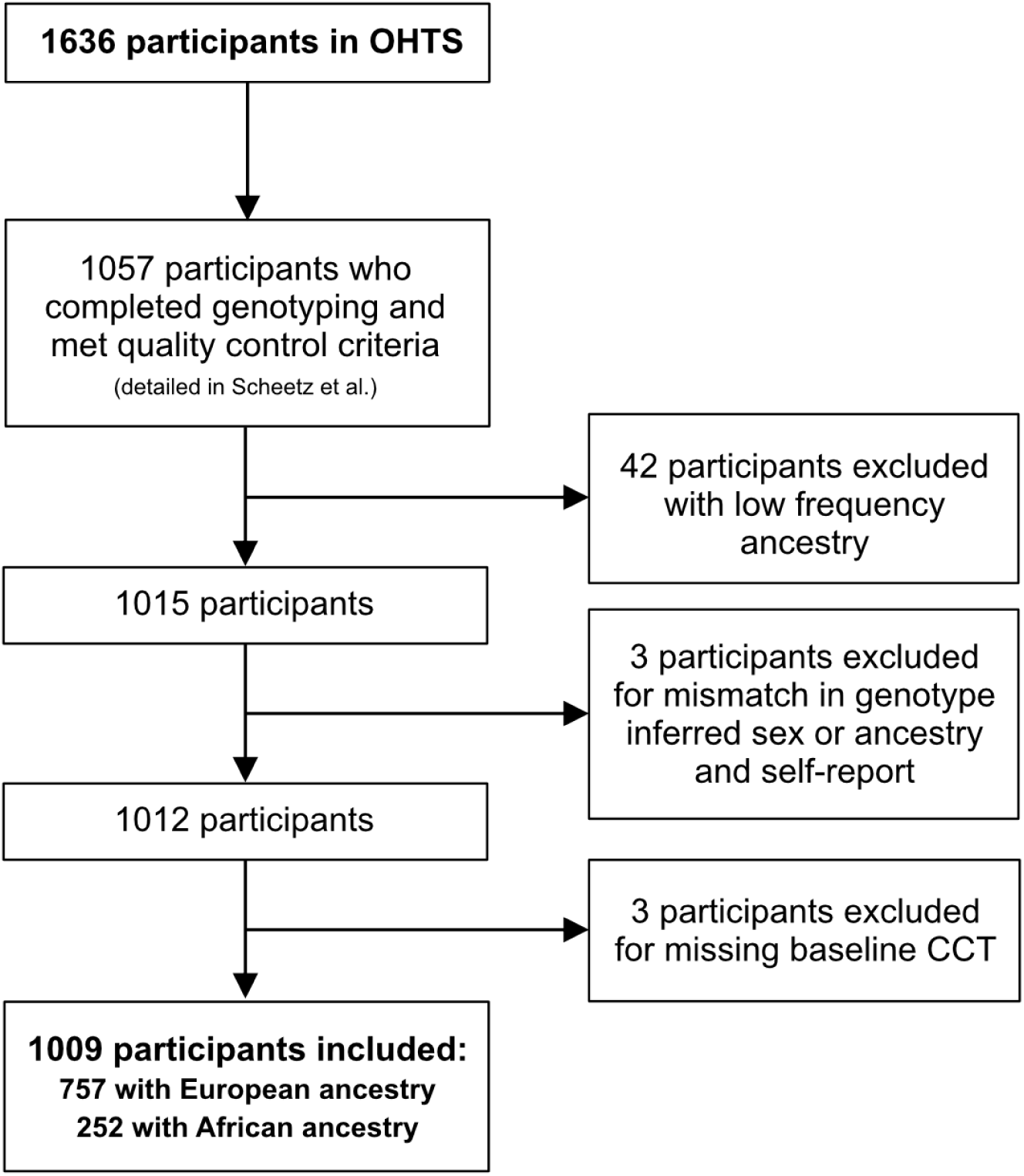
Participant flow chart describing inclusion and exclusion criteria of OHTS participants.

### Generation of polygenic risk score

We constructed a POAG PRS using summary statistics from the largest cross-ancestry POAG GWAS meta-analysis to date^10^ after exclusion of the UK Biobank (UKBB) cohort (summary statistics available at https://www.asegrelab.org/data). We trained our PRS within the UKBB population using the Lassosum penalized regression framework method with the EUR.hg19 1000 Genomes Project reference panel to inform LD structure (Fig 2A and 2B)^10,30^. The PRS was trained using 8,813,496 genomic variants from 449,186 cross-ancestry participants in the UK Biobank (UKBB) (Fig 2B). Prior to training the PRS, we applied various genotype array quality control steps with PLINK 1.9/PLINK 2 on directly genotyped and imputed variants of 449,186 UKBB samples similar to by Bycroft et al^31^ as described in detail in Supplement 2. ^10,30^

**Figure 2.**
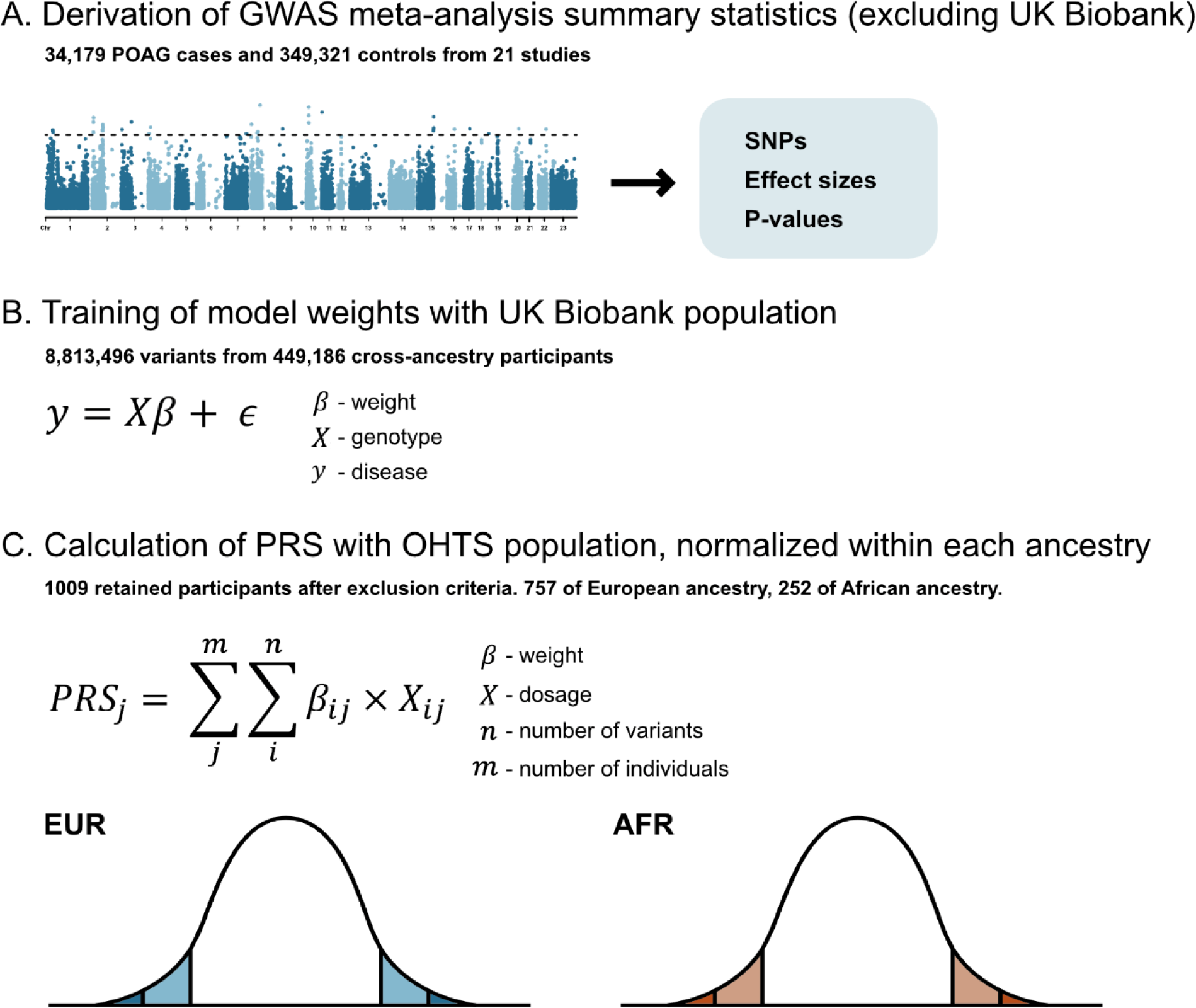
Description of the three-stage process for generation of a PRS in OHTS participants. A. Derivation of GWAS meta-analysis summary statistics. Example Manhattan is plot shown. B. Training of PRS in cross-ancestry UKBB population. C. Calculation of PRS in OHTS population.

The calculated weights from the UKBB training were then used to compute PRS using the PLINK ‘--score’ function among 1009 OHTS participants while retaining dosage information for imputed variants (Fig 2C). Specifically, our OHTS PRS included 5,235,564 variants that overlapped between the OHTS and UKBB genotype arrays. Lastly, as PRS is a relative measure within a population^13^, we transformed our PRS to be centered at 0 with standard deviation of 1 (z-score), within each inferred ancestry, as recommended^32–34^. We did not use raw PRS values derived from UKBB-trained weights directly with the OHTS participants, as raw OHTS PRS was much higher than that of the UKBB participants, even after selection for high IOP (Supplement 3 Figure 1). Within OHTS participants, mean unadjusted PRS scores were significantly higher for those of African ancestry compared those of European ancestry (Supplement 4 Fig 1), a phenomenon that has been observed in other studies and is likely due to differences in the sizes of linkage disequilibrium blocks and variant allele frequencies among ancestral backgrounds^33^. To correct for this, PRS values were transformed to Z-scores within each of inferred European (N=757) and African ancestries (N=252) (Fig 2C). An alternate set of results using raw PRS without transformation is provided in Supplement 5 to demonstrate the robustness of our results.

### Baseline prediction models

Survival regression analysis was performed to predict time to the event of interest: conversion of OHTN to POAG. Cox-Proportional Hazards (Cox-PH) models were implemented with scikit-survival 0.19.0 and lifelines 0.27.4 libraries on a Python 3.9.12 platform^35,36^. Two multivariable Cox-PH models with the following input eye-level baseline covariates were generated: 1) Model 1: age, IOP, PSD, VCDR, CCT, randomization status (RS), and 2.) Model 1 plus PRS. RS was included in all models to control for effect of delayed treatment on POAG onset in OHTS phase 1 participants^29^. Lastly, to determine the predictive performance of each baseline covariate individually, simplified univariate models for each variable of interest were constructed, with adjustment for RS in each model.

### Statistical analysis

Mean normalized PRS for each European and African ancestry was compared with a two-sample t-test, and for differences by baseline risk tertile as defined by OHTS prediction model with one-way analysis-of-variance (ANOVA). Participants were then grouped into either deciles or low, intermediate, and high risk tertiles based on PRS Z-score. Differences in participant-level demographics, follow-up duration, and baseline ocular parameters were assessed with one-way ANOVA. Differences in POAG-onset over 20 years between risk tertiles were compared with K-sample log-rank test. The relationship between age-at-diagnosis and PRS decile was assessed with linear regression adjusted for ancestry and gender, with and without stratification for early treatment status.

For the prediction of 20-year POAG onset, data was divided into training-testing sets with 90- 10% splits, and ten-fold cross validation (CV) was performed. Group k-fold methodology was applied so both eyes from the same patient appeared in the assigned training or testing sets. Concordance-index was computed over each CV fold to determine overall predictive performance and the mean score is provided for each model. Cumulative/dynamic ROC-AUC (AUC(t)) scores were computed to assess model stability, as sensitivity and specificity of survival regression models vary with time^37–39^. AUC(t) was calculated in 0.5-year intervals from 2.25 years to 20 years post-randomization. This period was chosen because randomly generated CV folds often exclude participants with conversion before 2.25 years. Mean AUC(t) was calculated for OHTS phases 1, 2, and 3. The predictive performance of models 1 and 2 was compared using Wilcoxon signed-rank test with mean cross-validation AUC(t) scores.

For all Cox-PH models, hazard ratios (HR) were calculated with the entire data set after input variables were scaled to fit normal distributions centered at 0 with a standard deviation of 1. HR represents a single standard deviation increase in continuous variables and change of state in binary variables.

## Results

Mean PRS was significantly higher for POAG-converters (0.24 ± 0.95) than for non-converters (- 0.12 ± 1.00) (p < 0.01). When stratified by ancestry, mean PRS for POAG-converters of European descent (0.29 ± 0.95) was significantly higher than corresponding non-converters (-0.13 ± 0.99) (p<0.01). For participants with African ancestry, mean PRS was higher for POAG-converters (0.13 ± 0.93) than for non-converters (-0.10 ± 1.04), but did not reach statistical significance (p = 0.07). There was a significant difference in PRS between low (-0.08 ± 1.03) and high (0.18 ± 0.94) baseline risk tertiles (p = 0.05) as defined by the original OHTS prediction model^8^. When stratified by European ancestry, there was no significant difference in PRS between low (-0.07 ± 0.98) and high (0.10 ± 1.01) baseline risk tertiles (p=0.31). For participants with African ancestry, the difference in PRS between low (-0.14 ± 1.26) and high (0.27 ± 0.85) baseline risk tertiles was not statistically significant (p=0.10).

When PRS is aggregated into risk tertiles, there were no significant differences in baseline age, gender, IOP, or CCT (Table 1). There was a significant decrease in follow-up duration and increase in VCD with increase in risk tertile (Table 1).

**Table 1.** Description of entire study population and comparison of baseline characteristics in PRS risk tertiles. ** represents significance threshold of 0.05 for ANOVA test between PRS risk tertiles.

|  |  | PRS Risk Tertile |  |  |  |
| --- | --- | --- | --- | --- | --- |
|  | Study population (n=1009) | Low (n=337) | Intermediate (n=323) | High (n=349) | p |
| Age (y) | 55.93 | 56.29 | 55.73 | 55.76 | 0.68 |
| Gender (% Male) | 44.30% | 43.91% | 43.96% | 45.00% | 0.95 |
| Follow-up duration (y) | 15.20 | 15.82 | 14.97 | 14.81 | **0.03\*\*** |
| IOP (mmHg) | 24.94 | 24.72 | 24.89 | 25.2 | 0.07 |
| VCDR | 0.39 | 0.36 | 0.38 | 0.43 | **<0.01\*\*** |
| CCT (um) | 573.43 | 574.24 | 572.62 | 573.39 | 0.86 |
| PSD (dB) | 1.91 | 1.91 | 1.91 | 1.91 | 0.97 |

There was significantly increased risk for POAG onset in the highest tertile of PRS compared to those in the lowest tertile, by the end of OHTS phase 1 (3.01-fold increase in POAG-onset, p<0.01), phase 2 (2.30-fold increase, p<0.01), and phase 3 (2.00-fold increase, p<0.01) (Fig 3A). When stratified by ancestry, there was a significant difference in POAG-free survival between low and high PRS tertiles for European descent participants by the end of OHTS phase 1 (4.37-fold increase, p<0.01), phase 2 (2.37-fold increase, p<0.01), and phase 3 (2.00-fold increase, p<0.01) (Fig 3B). For African ancestry participants, there was a significant difference between high and low PRS tertiles by the end of OHTS phase 2 (2.14-fold increase, p<0.01) and phase 3 (2.04-fold increase, p<0.01), but not by the end of phase 1 (p=0.18) (Fig 3C).

**Figure 3.**
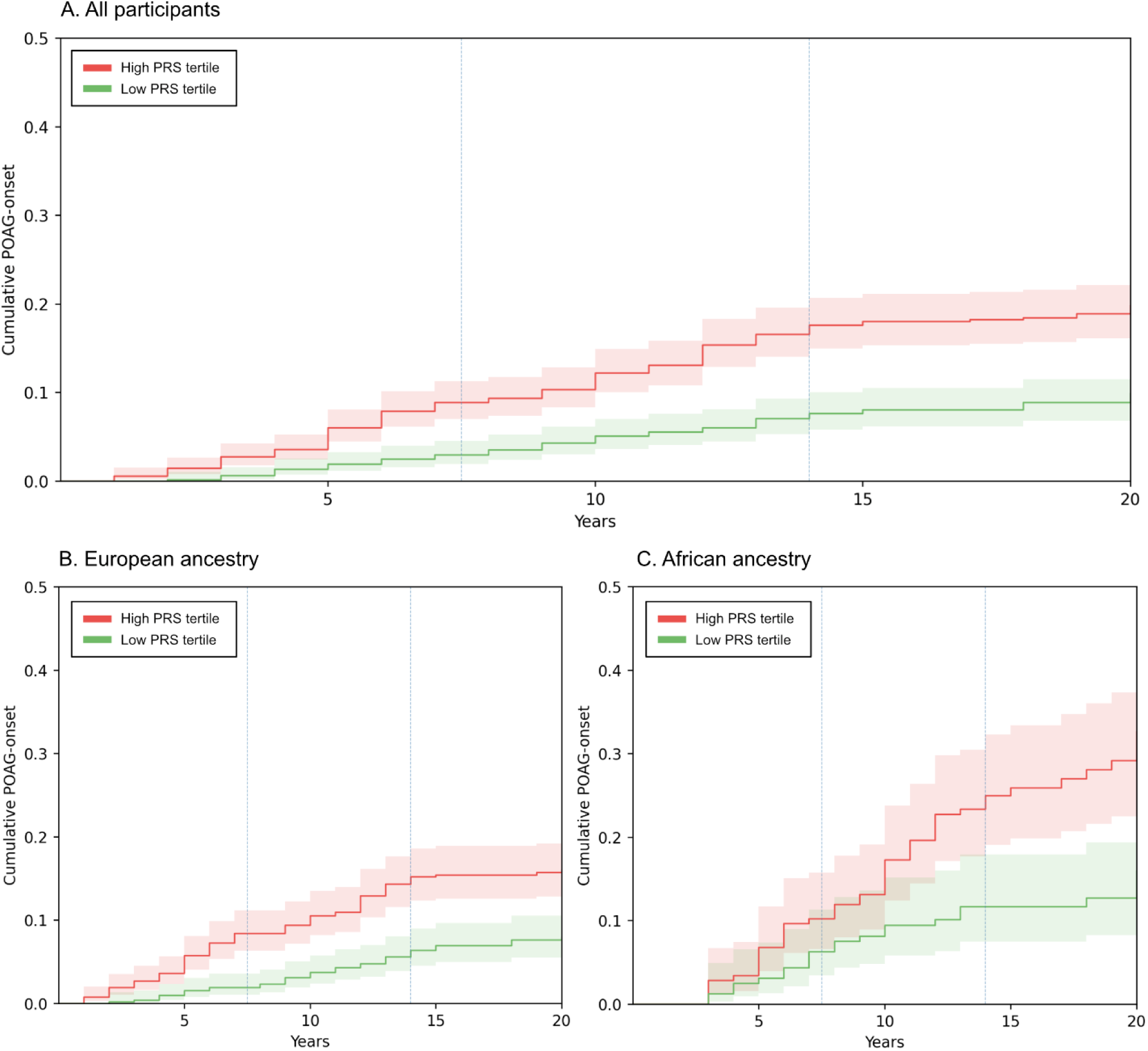
Cumulative POAG-onset curves, adjusted for exposure time, for A. All participants, B. European ancestry, C. African ancestry.

Each decile of higher PRS was associated with 2.46% (CI=[2.10-2.81%]) greater POAG-conversion risk per eye, with linear regression showing an increase from 14.85% conversion over 20 years in lowest decile to 36.99% conversion at highest decile, in analysis without adjustment for exposure time (Fig 4A). With adjustment for exposure time, each decile in PRS was associated with a 1.36% (CI=[1.08-1.64%])greater POAG-conversion risk, with linear regression showing an increase from 9.52% 20-year conversion in the lowest PRS decile to 21.81% in the highest PRS decile (Fig 4B).

**Figure 4.**
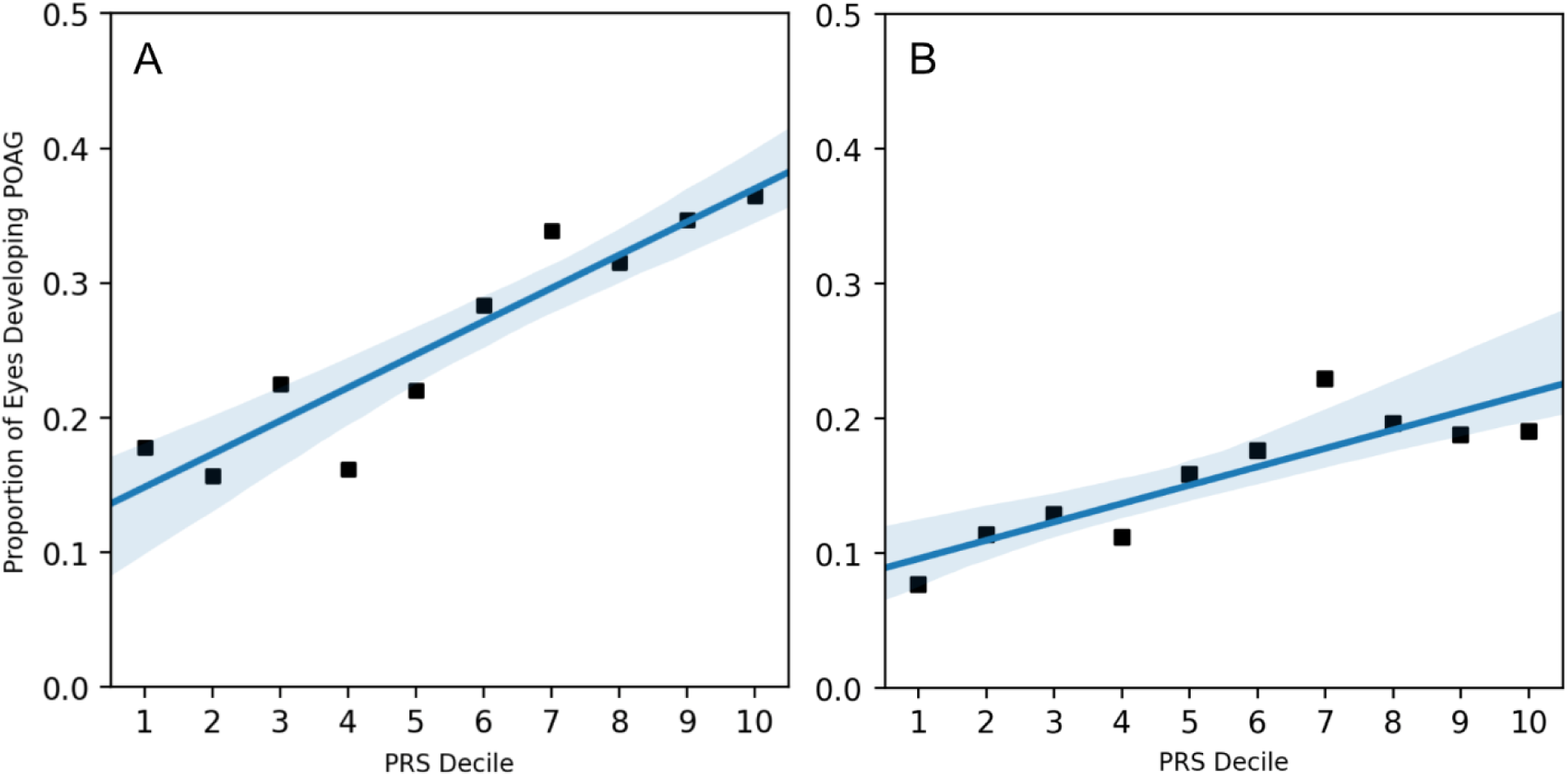
The proportion of participants developing POAG by the end of OHTS phase 3 (20 years) plotted against PRS decile. A. Without adjustment for exposure time. B. With adjustment for exposure time. Fitted linear regression is also shown, with 95% confidence interval around mean prediction.

In multivariable linear regression with age at POAG diagnosis against PRS decile, no significant linear relationship is initially found (beta = 0.34 years younger age at diagnosis per higher decile, P=0.058) (Fig 5A). When stratified by delay in OHTN treatment, those with delayed treatment had a significant linear relationship between age at diagnosis and PRS decile (beta = 0.52 years younger age at diagnosis per higher PRS decile, P=0.047). Those who received early treatment had no significant relationship (beta = 0.17 years earlier diagnosis per higher decile, P=0.493). (Fig 5B-C).

**Figure 5.**
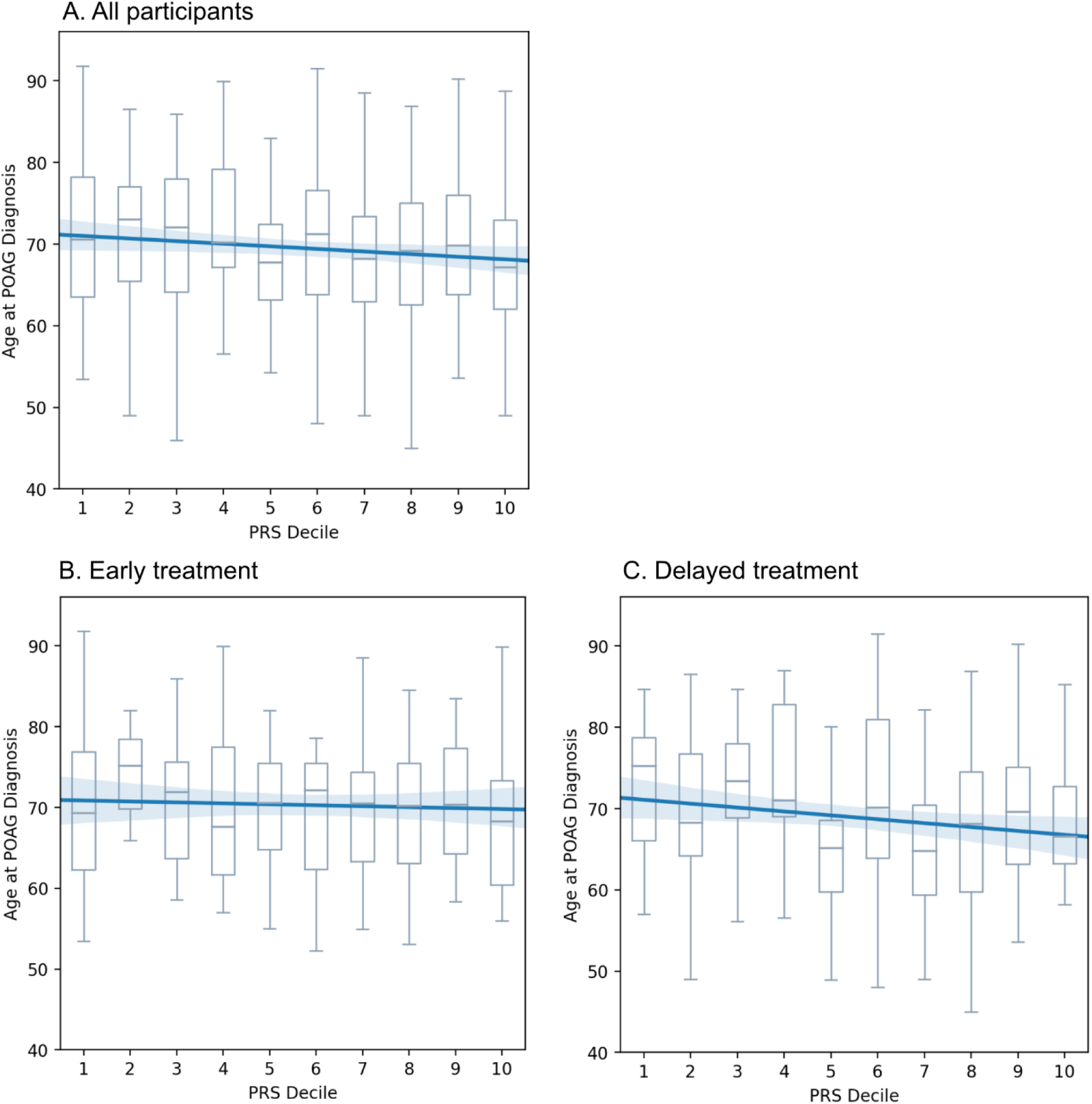
Age at diagnosis, with 95% confidence interval, plotted against PRS decile. Fitted univariate linear regression also shown, with 95% confidence interval around mean prediction (shaded area).

AUC(t) for multivariable Cox-PH prediction models is shown in Fig 6. Average performance over each phase of OHTS and overall C-index are shown in Table 2A. Model 2, which added PRS improved mean AUC(t) in OHTS phases 1, 2, and 3 as well as overall C-index over Model 1 (p<0.01). In analysis limited to European ancestry participants, C-index for Model 2 is 0.78 and C-index for Model 1 is 0.76 (p<0.01). In participants with African ancestry, C-index for Model 1 is 0.70 and for Model 2 is 0.71 (p=0.21).

**Figure 6.**
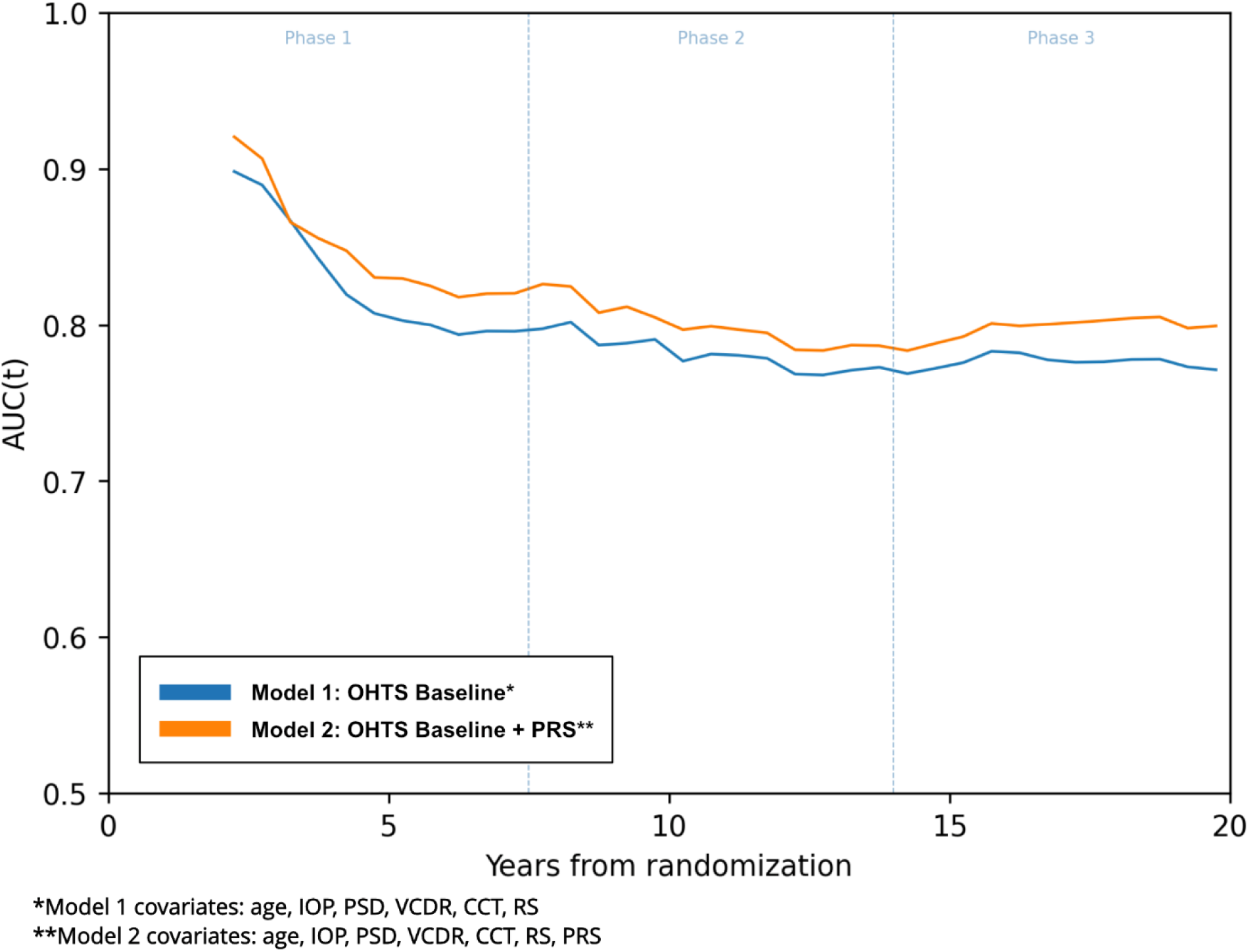
AUC(t) curves for baseline prediction models, calculated in 0.5 year intervals.

**Table 2.** Comparison of baseline prediction models. Average AUC(t) is shown within each phase of OHTS as a proxy for early, intermediate, and late conversion to POAG. A. Multivariable models. B. Univariate models for continuous variables, adjusted for randomization status (RS).

| **2A. Multivariable prediction models** |  |  |  |  |
| --- | --- | --- | --- | --- |
| **Model** | **Average AUC(t)**<br>**(Phase 1)** | **Average AUC(t)**<br>**(Phase 2)** | **Average AUC(t)**<br>**(Phase 3)** | **C-Index** |
| Model 1* | 0.83 | 0.78 | 0.77 | 0.75 |
| Model 2** | 0.85 | 0.81 | 0.79 | 0.77 |
| **2B. Simplified prediction models, only adjusted for randomization status** |  |  |  |  |
| **Model** | **Average AUC(t)**<br>**(Phase 1)** | **Average AUC(t)**<br>**(Phase 2)** | **Average AUC(t)**<br>**(Phase 3)** | **C-Index** |
| VCDR | 0.72 | 0.70 | 0.69 | 0.68 |
| CCT | 0.71 | 0.68 | 0.64 | 0.63 |
| IOP | 0.70 | 0.67 | 0.63 | 0.63 |
| PRS | 0.67 | 0.67 | 0.63 | 0.63 |
| Age | 0.66 | 0.65 | 0.65 | 0.62 |
| PSD | 0.68 | 0.64 | 0.61 | 0.61 |

\*Model 1 covariates: age, IOP, PSD, VCDR, CCT, RS
\*\*Model 2 covariates: age, IOP, PSD, VCDR, CCT, RS, ancestry, PRS

A one standard deviation higher PRS conferred a hazard ratio of 1.25 (CI=[1.15-1.37]) for conversion to POAG in Model 2. Simplified models to assess predictive performance of each baseline covariate demonstrated that VCDR was the best predictor overall based on C-index. (Table 2B).

## Discussion

We show that a high POAG PRS, based on the largest GWAS metanalysis to date, was associated with increased risk of POAG conversion among patients with ocular hypertension. This effect was independent of most phenotypic baseline risk factors and was independent of stratification by patient ancestry. Furthermore, it holds in both the early and late phases of the OHTS follow-up period. We also found that the cumulation of background genetic risk is associated with younger age at diagnosis in participants who received delayed treatment. Lastly, we demonstrate that the inclusion of a PRS improves multivariable risk stratification models.

PRS was a strong predictor of POAG risk in our OHTN study population, with the risk of conversion increasing 2.3-fold from the lowest PRS decile to the highest, and significantly increased 20- year POAG incidence in participants of European and African descent. When PRS is grouped into risk tertiles, the incidence of conversion is 3.0-fold higher in early conversion (OHTS phase 1) and decreases to 2.0-fold in late conversion (OHTS phase 3). These increases in incidence are higher than the 12% increase in previous attributed to TMCO1 locus risk alleles in OHTS^27^.

When stratified by ancestry, this relationship held for European ancestry over all phases of OHTS, and African ancestry over OHTS phases 2 and 3. Comparison of low and high PRS risk tertiles in those with African ancestry is likely underpowered in OHTS phase 1, due to the low number who developed POAG. A decrease in risk attributable to PRS tertile in later phases of OHTS, is most likely due to an increasing contribution of age or cumulative environmental burden. While the increase in absolute risk appears lower than the 6 to 45-fold increases seen in other studies, it is important to note that our study includes only OHTN patients with healthy optic nerves at baseline, but increased risk of conversion compared to those with normal IOP^20,40,41^. Furthermore, OHTN patients who do not develop glaucoma may have undiscovered protective variants not captured by our PRS.

Analysis of collinearity between PRS and other baseline covariates only showed a significant relationship with VCDR, with a modest increase from 0.36 to 0.43 from lowest to highest PRS tertile (Table 1). This suggests that genetic variants influencing VCDR may also play a role in the conversion of OHTN to POAG. While prior work has shown certain risk variants to be associated with IOP, we did not find a statistically significant difference in IOP between the lowest PRS to highest PRS tertiles (Table 1, p=0.07)^10,42,43^. It is possible that IOP association effects are limited by selection bias for high IOP at baseline. A clinically important finding is the loss of significance of the association between PRS and age at diagnosis for participants receiving early treatment. This suggests that the onset of clinically detectable disease was delayed by 5.2 years with early treatment. This relationship is potentially understated in our analysis, as nearly half of POAG non-converters were lost to follow-up and high age at enrollment was correlated with attrition.

Importantly, Cox-PH prediction models demonstrated improvement in the prediction of POAG-onset with addition of PRS. A one-standard-deviation higher PRS increased the 20-year risk for POAG by 25%. This is consistent with our findings of relative independence of PRS from other baseline covariates. PRS added a relatively constant improvement in prediction over time, with AUC(t) improvements in both early-and late-phases of OHTS, most likely because it is a measure of constant, genetically determined risk. The absolute increase in performance of prediction models, from C-index of 0.75 to 0.77, may be smaller than expected given the independence of PRS from most baseline covariates. This may be due to selection bias in this population, as OHTN alone confers increased risk of POAG, and the aggregate phenotype represented by VCD, CCT, and PSD may share relationships with PRS not represented by univariate collinearity analysis alone. Furthermore, the absence of newer phenotypic baseline information, such as structural optical coherence tomography, may be limiting potential improvement in predictive models. Future studies incorporating modern imaging modalities should consider replication of our findings with PRS in newer risk stratification models.

Simplified predictive models showing prediction from each baseline covariate alone, demonstrated that VCDR was the strongest predictor of POAG conversion over the entire tested follow-up period. PRS was a stable predictor, showing an AUC(t) of 0.67 in early phases of OHTS and 0.63 in phase 3. CCT, baseline IOP, and PSD were most predictive of early POAG-onset and showed decreased in AUC(t) over time. Age was a stable predictor that gains relative importance over time, becoming the second most important predictor during OHTS phase 3. This is consistent with our findings showing a relative decrease in decreased risk stratification by PRS tertile during later phases of OHTS.

A remaining challenge in the incorporation of PRS in clinical care is susceptibility to bias from how well individual patients are represented by the study population^12^. We normalize PRS within OHTS participants, by ancestry, in this study but found differences in OHTS PRS distribution compared to the study population in the UKBB (Supplement 3 Fig 1). Furthermore, addition of PRS to the OHTS model does not significantly improve performance when analysis is limited to African ancestry participants. The comparison of mean PRS for POAG-converters versus non-converters also does not reach significance in the African ancestry subset. While these comparisons may be underpowered due to much lower sample size, other possibilities include limitations of the PRS itself. Most participants in prior POAG GWAS are of European descent and thus prior GWAS may be underpowered for identification of risk alleles associated with other ancestries. While moderate-to-high correlation in risk loci effect sizes has been shown between European, African, and Asian ancestries, new ancestry-specific variants may be discovered with GWAS targeting respective populations^10^ therefore allowing for improved, ancestry specific PRS development. Moreover, PRS may only represent a portion of genetically determined risk for disease as it only includes single nucleotide polymorphism-based heritability with a frequency of greater than 1% in the study population^44,45^.

The strengths of our study include those of the prospective OHTS data set; 20 years of data from close follow-up in a diverse sample, with validation of POAG onset by reading centers and an endpoint committee. Therefore, our results are less likely to be biased by delay in onset and formal diagnosis in asymptomatic stages. In addition, PRS were generated from a very large sample with the greatest number of alleles identified to date. Limitations include the relatively smaller size of the OHTS cohort, and low proportion of participants with confirmed POAG likely limited the statistical power of our stratified analyses. This and the loss to regular follow-up 15-20 years post-randomization may impact the predictive performance of our models. Furthermore, while we perform cross-validation to improve internal validity, validation of a PRS from the OHTS population in an external study population would improve generalizability of our results.

In conclusion, high polygenic risk for POAG is associated with increased risk for and earlier development of POAG in patients with OHTN, independent of most ocular phenotypic risk factors, patient ancestry, and duration of follow-up up to 20 years. Early treatment may mitigate risk from high genetic burden, delaying clinically detectable disease by up to 5.2 years.

## Data Availability

Genomic data from OHTS is available online through the National Eye Institute. The remainder of the OHTS data set will be made available at the NEI Data Commons.

https://www.ncbi.nlm.nih.gov/projects/gap/cgi-bin/study.cgi?study_id=phs000240.v1.p1

https://ohts.wustl.edu/results/

## Notes

**Conflict of interest:** Dr. Pasquale is a consultant for Twenty Twenty and Character Biosciences

### Competing Interest Statement

The authors have declared no competing interest.

### Clinical Trial

NCT00000125

### Clinical Protocols

https://ohts.wustl.edu/results/

### Funding Statement

- NEI K23EY032634
- NEI R01EY032559
- Research to Prevent Blindness Career Development Award
- American Glaucoma Society Clinician Scientist Award
- The Glaucoma Foundation (NYC)

### Author Declarations

Ethics committee/IRB of Mass General Brigham name waived ethical approval for this work

