## Supplemental Appendix 1 for "Polygenic Risk Score Improves Prediction of Primary Open Angle Glaucoma Onset in the Ocular Hypertension Treatment Study"

### 1. Derivation of ancestry in the Ocular Hypertension Treatment Study (OHTS) population

For the derivation of ancestry in OHTS population, Principal Component Analysis (PCA) was first applied to linkage disequilibrium (LD)-pruned ($r^2$<0.2 in 200kb windows) genetic markers with minor allele frequency (MAF)>5%. The SHARDS algorithm selected the first 3 important PCs and then implemented k-nearest neighbors classification to predict the ancestral background of participants using labels from the 1000 Genomes Project Phase 3 reference panel (Fig S1). After that, participants were categorized based on primary genetic ancestry into European (EUR), African (AFR), East Asian (EAS), South Asian (SAS), and Admixed American (AMR) descent (Table S1). Due to the small sample size, participants with inferred EAS, SAS, and AMR or self-reported Asian, admixed, or Hispanic descent were excluded (Table 1). European and African ancestry were most common in OHTS participants and retained for this analysis. Two samples were removed for mismatch between self-reported European and inferred AFR ancestry (n=1012). European population was the sum of self-report and inferred EUR without mismatch (n=760). African population was the sum of self-report and inferred AFR without mismatch (n=252).

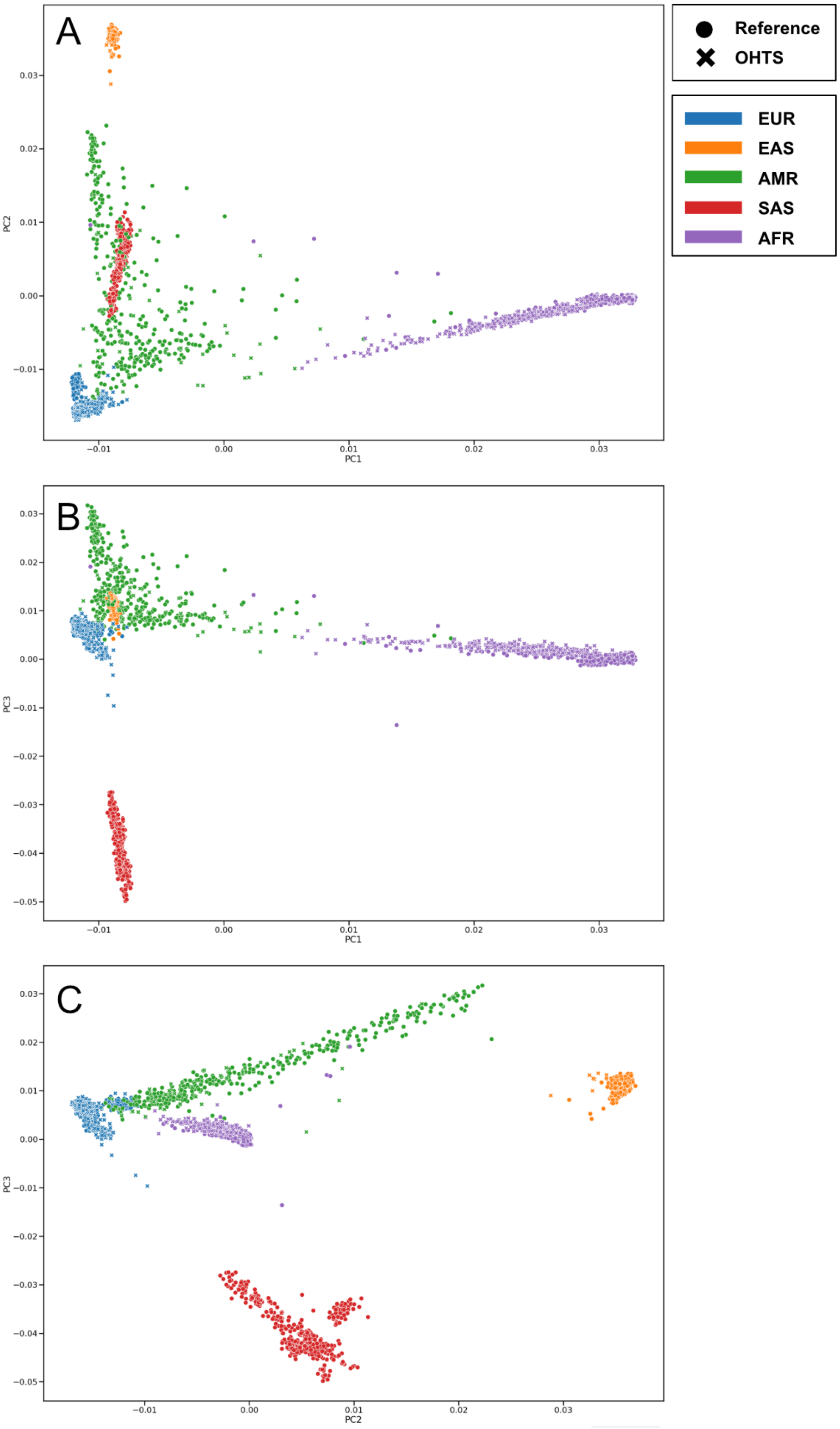


**Fig 1.** Ancestry classification plots for top 3 principal components (PC1, PC2, PC3, respectively) via SHARDS algorithm A. PC2 vs. PC1. B. PC3 vs PC1. C. PC3 vs PC2.

| | Predicted EUR | Predicted AFR | Predicted SAS | Predicted EAS | Predicted AMR |
|---|---|---|---|---|---|
| **Reported European** | 744 | 2** | 0 | 0 | 5 |
| **Reported African** | 0 | 246 | 0 | 0 | 3 |
| **Reported Asian** | 0 | 0 | 1** | 7** | 1** |
| **Reported Admixed** | 1 | 0 | 0** | 0** | 1** |
| **Reported Hispanic** | 8 | 2 | 0** | 1** | 25** |
| **Reported Other** | 2 | 1 | 1** | 0** | 5** |

**Table 1.** Self-reported race tabulated against inferred ancestry in OHTS population. ** denotes participants excluded from analysis in this study.
