## Supplemental Appendix 2 for "Polygenic Risk Score Improves Prediction of Primary Open Angle Glaucoma Onset in the Ocular Hypertension Treatment Study"

### 2. UK Biobank Quality Control

For samples with mismatched ancestry, we utilized the inferred ancestry for downstream analyses. Hard call sample quality control (QC) was conducted to remove withdrawn samples (N=107), samples not included in the imputed dataset (N=969), samples with no sample QC data (N=112), ambiguous sex (N=1930), heterozygosity outliers by using 5 standard deviations from the mean heterozygosity rate within each inferred ancestry subpopulation (N=978) and individuals with high cryptic relatedness (>0.1875 Pi-hat) (N=35,113). Graph theory was applied to keep the maximal independent set and the highest ratio of cases versus controls to improve the power. Next, imputed variant QC was performed using a pipeline that iteratively examines variants (single nucleotide polymorphisms [SNPs] and indels [insertions and deletions]), INFO score, allele frequencies and tests of Hardy-Weinberg equilibrium. More specifically, the missingness rate per sample using posterior probability>0.9 for genotype calling was inspected. A small missingness rate of autosomes was observed and no significant difference was observed between male and female. Monomorphic sites (M=552,024) were removed if only one allele occurs at a site or locus in the population. The monomorphic cutoff was defined as

$$Monomorphic\ cutoff = \frac{1}{2.1 * N}$$

where N is the number of samples. We further filtered out the variants with indels length>50 bp (M=2,312) and subsequently, variants with INFO score<0.6 (M=45,927,754). The variants with allele frequency<0.1% for each ancestry subpopulation (EUR: M=35,850,305; AMR: M=28,792,905; AFR: M=24,954,341; SAS: M=35,437,737; EAS: M=37,236,001) were further excluded. Hardy-Weinberg equilibrium was calculated and inspected for each ancestry subpopulation. The mid p-value was utilized in exact tests for Hardy-Weinberg equilibrium analyses[42]. Therefore, a total of 227,173 variants in EUR, 98 variants in AMR, 32,317 variants in AFR, 38,173 variants in SAS, and 3,711 variants in EAS of autosomes failed Hardy-Weinberg (P<1e-06) and were removed. To be conservative, a variant was kept if it passed the Hardy-Weinberg for all ancestry subpopulations. Finally, a mean allele frequency (MAF) filter was applied to retain the common variants with MAF>1% for further analyses.
