## Supplemental Appendix 3 for "Polygenic Risk Score Improves Prediction of Primary Open Angle Glaucoma Onset in the Ocular Hypertension Treatment Study"

### 3. Comparison of Ocular Hypertension Treatment Study (OHTS) and UK Biobank (UKBB) polygenic risk Score (PRS)

OHTS participants have higher PRS than UKBB participants, even after selection for high IOP within the UKBB (Figure S3).

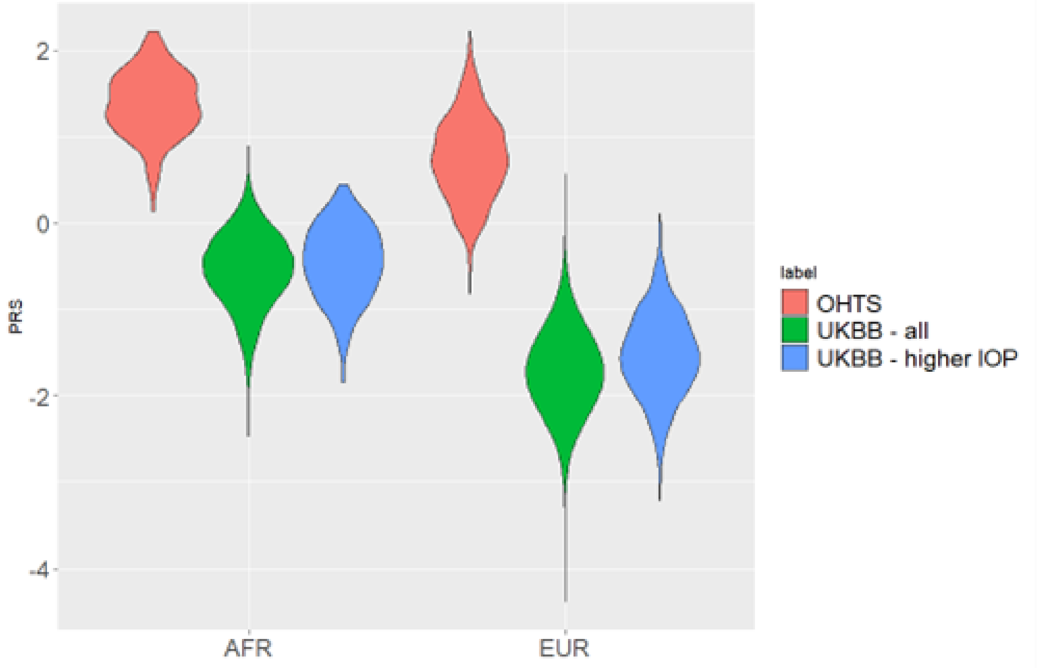


**Fig 1.** Violin plots comparison PRS distributions in participants from OHTS, UKBB, and UKBB selected for high IOP (corresponding to OHTS inclusion criteria).
