## Supplemental Appendix 4 for "Polygenic Risk Score Improves Prediction of Primary Open Angle Glaucoma Onset in the Ocular Hypertension Treatment Study"

### 4. Polygenic risk score (PRS) distributions by ancestry

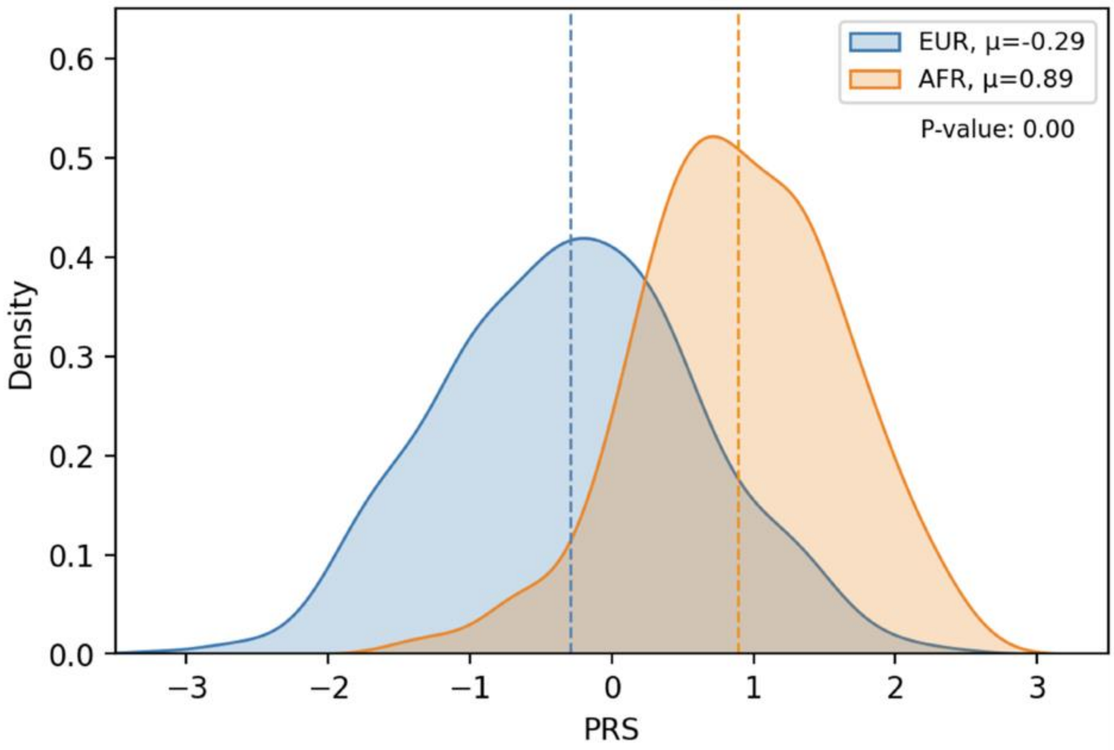


**Fig 1.** Density plot of PRS distributions by ancestry, normalized to entire population. The EUR curve represents European ancestry. The AFR curve represents African ancestry.
