## Supplemental Appendix 5 for "Polygenic Risk Score Improves Prediction of Primary Open Angle Glaucoma Onset in the Ocular Hypertension Treatment Study"

**5. Results with raw polygenic risk score (PRS). In this analysis, there is no transformation within inferred ancestries.**

A. Raw PRS distributions in OHTS participants of European ancestry

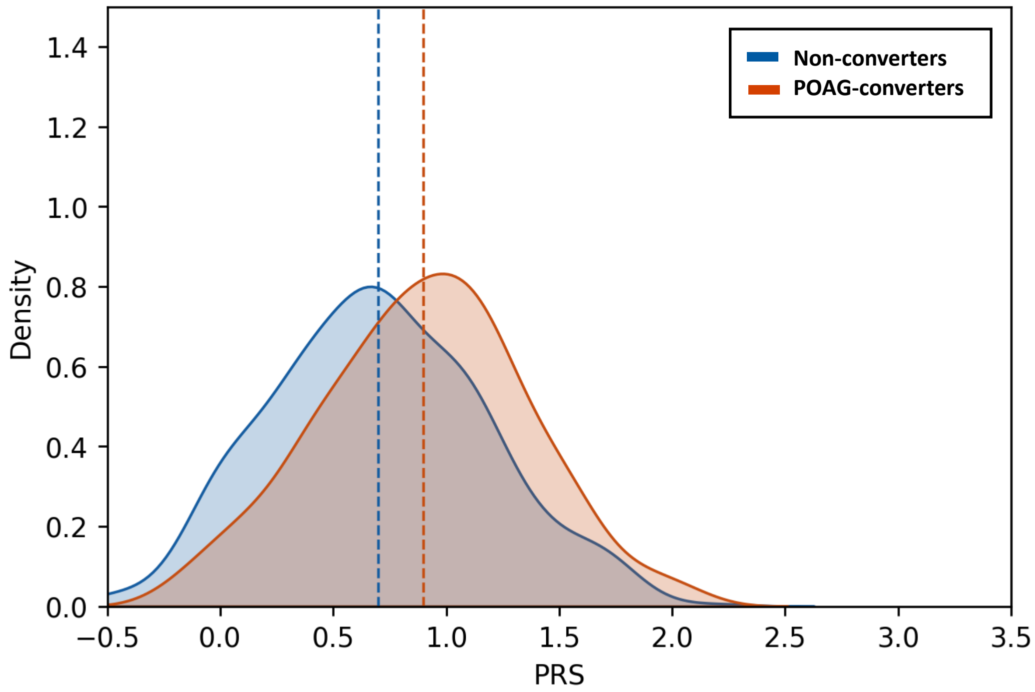


B. Raw PRS distributions in OHTS participants of African ancestry

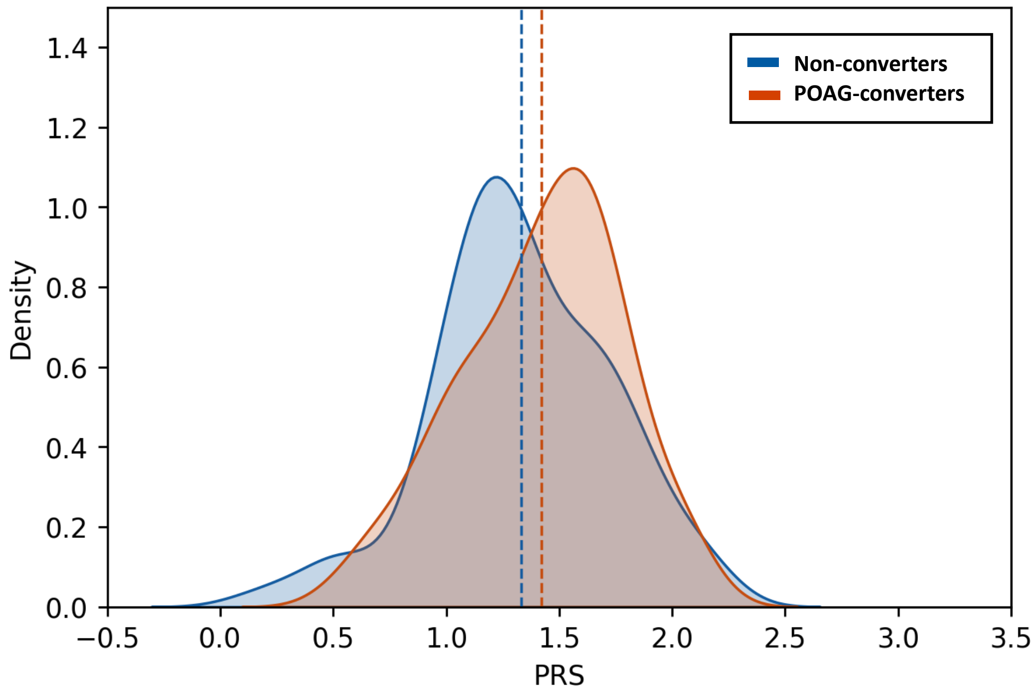

**Figure 1.** Distributions of PRS compared between POAG-converters (red) and non-converters (blue). A. Shows distribution in participants with European ancestry. B. Shows distributions in participants with African ancestry.

AUC(t) for multivariable Cox-PH prediction models is shown in Fig 2. This adds a third model using raw PRS to supplement Figure 6 in the primary results.

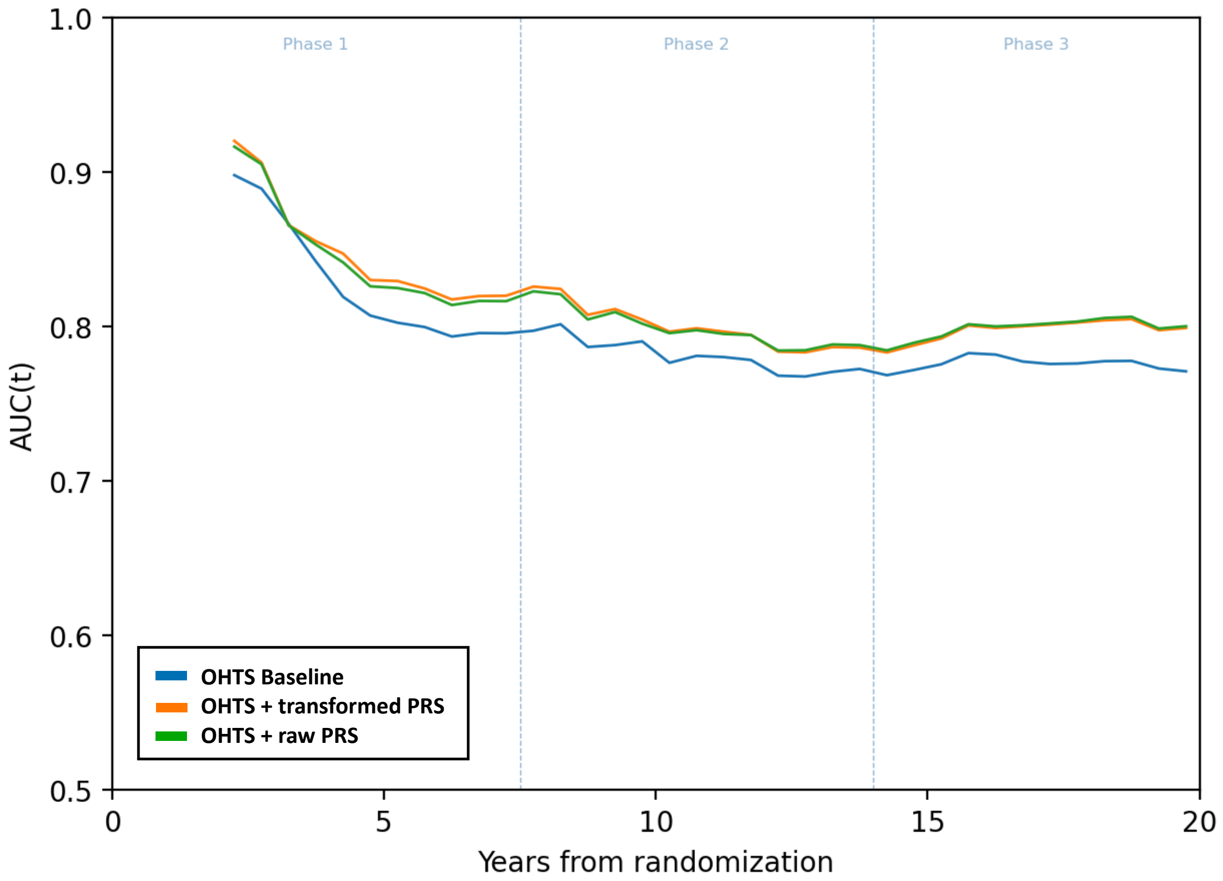


**Figure 2.** AUC(t) curves for baseline prediction models, calculated in 0.5-year intervals. A third model with raw PRS without transformation by ancestry is included here.

In analysis limited to European ancestry participants, C-index for Model 2 is 0.78, and C-index for Model 1 is 0.76 ($p<0.01$). In participants with African ancestry, C-index for Model 1 is 0.70 and for Model 2 is 0.71 ($p=0.21$). There are no meaningful differences in model performance using raw PRS or scaling to a Z-score within each genetically-derived ancestry. However, scaling eases the interpretation of effect sizes relative to a reference population compared to raw PRS.
